# Staff testing for COVID-19 via an online pre-registration form

**DOI:** 10.1101/2020.07.13.20152876

**Authors:** M. Saadiq Moolla, Arifa Parker, M. Aslam Parker, Sthembiso Sithole, Leila Amien, Rubeena Chiecktey, Tasneem Bawa, Abdurasiet Mowlana

## Abstract

**Background:** Healthcare workers are at increased risk of contracting SARS-CoV-2 and potentially causing institutional outbreaks. Staff testing is critical in identifying and isolating infected individuals while also reducing unnecessary workforce depletion. Tygerberg Hospital implemented an online pre-registration system to expedite staff and cluster testing.

**Objectives:** We aimed to identify (1) specific presentations associated with a positive or negative result for SARS-CoV-2 and (2) staff sectors where enhanced strategies for testing might be required.

**Methods:** Retrospective descriptive study involving all clients making use of the hospital’s pre-registration system during May 2020.

**Results:** Of 799 clients, most were young and female with few comorbidities. The most common occupation was nurses followed by administrative staff, doctors and general assistants. Doctors tested earlier compared to other staff (median: 1.5 vs 4 days). The most frequent presenting symptoms were headache, sore throat, cough and myalgia. Amongst those testing positive (n=105), fever, altered smell, altered taste sensation, chills and history of fever were the most common symptoms. Three or more symptoms was more predictive of a positive test, but 12/145 asymptomatic clients also tested positive.

**Conclusion:** Staff coronavirus testing using an online pre-registration form is a viable and acceptable strategy. While some presentations are less likely to be associated with SARS-CoV-2 infection, no symptom can completely exclude it. Staff testing should form part of a bundle of strategies to protect staff including wearing masks, regular hand washing, buddy screening, physical distancing, availability of PPE and special dispensation for COVID-19-related leave.

**Unstructured Abstract:** Hospital staff testing is critical to identify and isolate SARS-CoV-2 infected individuals while reducing unnecessary workforce depletion. Tygerberg Hospital implemented an online pre-registration system to expedite staff testing. In a retrospective descriptive study of all clients registering during May 2020 (N=799), we found rapid and sustained uptake of the system. The most frequent presenting symptoms were headache, sore throat, cough and myalgia. Amongst those testing positive (n=105), fever, altered smell, altered taste sensation and chills were the most likely symptoms, but 12/145 asymptomatic clients also tested positive. Staff testing should form part of a bundle of strategies to protect staff.

## Introduction

The emergence of severe acute respiratory syndrome coronavirus 2 (SARS-CoV-2) has placed severe pressure on an already overburdened healthcare system in South Africa. At Tygerberg Hospital (TBH), initially the designated coronavirus disease 2019 (COVID-19) hospital in the Western Cape, South Africa, mitigation strategies have included the establishment of separately staffed high risk COVID-19 areas with clear signage, guidelines on the use of Personal Protective Equipment (PPE), staff education and training, and the construction of new infrastructure for triage, testing and treatment with multi-departmental involvement.^[1]^

These measures aimed to meet the challenge of COVID-19 while protecting staff, but healthcare workers remain at increased risk of contracting SARS-CoV-2, potentially causing institutional outbreaks and spreading infection back to their communities.^[2]^ This can result in the closure of entire departments or hospitals as has been reported in the media. TBH has not been immune, with numerous confirmed positive cases and COVID-19-related deaths reported amongst staff.

Isolation of potentially SARS-CoV-2 infected healthcare workers is critical in preventing nosocomial spread. However, workforce depletion in the context of existing staff shortages weakens the fight against COVID-19. A balanced approach is required.^[3]^

The Occupational Health Department (OHD) developed a risk assessment tool to identify individuals at risk for severe disease and place staff in the most appropriate areas to work. A system was also implemented to screen and test staff members, both symptomatic or contacts of confirmed cases, that would allow for early return to work according to current national guidelines, mitigating against staff shortages while at the same time identifying and isolating infected individuals.^[4]^

The necessity of staff testing created additional workload for the Triage and Testing Centre (TATC) which, by late April 2020 was routinely processing over 200 tests per day. Smoothly processing staff members rapidly became more difficult as the number presenting increased. To minimize waiting time and decrease the risk of cross infection from traditional paper-based formats, an online pre-registration system for staff and clusters was implemented. The form (available at www.tbhcovid.co.za/register) was completed by prior to arrival and included all relevant screening information. Once submitted, it generated an email to the clerks in the TATC allowing a folder to be opened. On arrival, staff in the TATC could retrieve the folder and test the individual in as little as 10 minutes.

We aimed to determine reasons for testing and specific presentations associated with positive or negative results. This information could be used to refine future healthcare worker testing and isolation guidelines. We also aimed to identify specific sectors of staff where testing was delayed or deficient and for whom more targeted interventions may be required.

## Methods

We conducted a single-centre retrospective descriptive study involving all cases created via the pre-registration system from 1 to 31 May 2020. All cases were included as there were no exclusion criteria.

We collected self-reported information from all emails received over the study period including date of registration, demographic information, symptoms, contact history, comorbidities and employment information. This data was exported from the inbox and algorithmically extracted before being checked for completeness and coded for statistical analysis. At our institution reverse transcriptase polymerase chain reaction for SARS-CoV-2 is performed on nasopharyngeal specimens. Test results were obtained from the National Health Laboratory Service (NHLS) TrakCare Web Results Viewer.

Data analysis was performed using VassarStats (available at http://vassarstats.net). Statistical significance was set at p < 0.05 and a 95% confidence interval (95% CI) was used.

Ethics approval for this article was obtained from the Health Research Ethics Committee of Stellenbosch University (ref. no. N20/04/002_COVID-19).

## Results

This study included 799 cases. The mean (standard deviation (SD)) age of clients was 39.7 (10.9) years old and the majority (619, 77.47%) were female. Most clients were from TBH (703, 87.98%) and 349 (43.68%) were nurses. Amongst nurses, 326 (93.41%) were female. Other groups included 150 (18.77%) administrative staff, 63 (7.88%) doctors and 58 (7.26%) general assistants. Most clients were healthy with 560 (70.09%) recording no comorbidities. The most common comorbidities were asthma (69, 8.64%), diabetes mellitus (61, 7.63%) and obesity (50, 6.26%). Hypertension was excluded from reporting due to a recording error. Table 1 depicts the baseline characteristics.

**Table 1:** Baseline characteristics.

| Characteristic (N = 799) | Frequency | Proportion (95% CI) |
| --- | --- | --- |
| Sex: Female | 619 | 77.47 (74.45 ; 80.23) |
| Institution |  |  |
| - Tygerberg Hospital | 703 | 87.98 (85.54 ; 90.06) |
| - Other Medical | 32 | 4.01 (2.86 ; 5.60) |
| - Non-medical | 21 | 2.63 (1.73 ; 3.99) |
| - Unallocated | 43 | 5.38 (4.02 ; 7.24) |
| Role |  |  |
| - Nurse | 349 | 43.68 (40.28 ; 47.14) |
| - Administrative staff | 150 | 18.77 (16.21 ; 21.62) |
| - Doctor | 63 | 7.88 (6.21 ; 9.96) |
| - General assistant | 58 | 7.26 (5.66 ; 9.27) |
| - Porter | 18 | 2.25 (1.43 ; 3.53) |
| - Allied health | 10 | 1.25 (0.68 ; 2.29) |
| - Radiology | 8 | 1.00 (0.51 ; 1.96) |
| - Security | 4 | 0.50 (0.19 ; 1.28) |
| - Laboratory | 1 | 0.13 (0.01 ; 0.81) |
| - Other or unallocated | 138 | 17.27 (14.81 ; 20.05) |
| Co-morbidities |  |  |
| - Asthma | 69 | 8.64 (6.88 ; 10.79) |
| - Diabetes mellitus | 61 | 7.63 (5.99 ; 9.68) |
| - Obesity | 50 | 6.26 (4.78 ; 8.16) |
| - Cardiac disease | 39 | 4.88 (3.59 ; 6.60) |
| - Previous history of tuberculosis | 33 | 4.13 (2.96 ; 5.74) |
| - HIV positive | 17 | 2.13 (1.33 ; 3.38) |
| - On antiretrovirals | 23 | 2.88 (1.88 ; 4.28) |
| - Pregnant | 12 | 1.50 (0.86 ; 2.60) |
| - Chronic kidney disease | 9 | 1.13 (0.60 ; 2.13) |
| - Other lung diseases | 9 | 1.13 (0.60 ; 2.13) |
| - Chronic liver disease | 6 | 0.75 (0.34 ; 1.63) |
| - Current tuberculosis | 4 | 0.50 (0.19 ; 1.28) |
| - None | 560 | 70.09 (66.83 ; 73.16) |

The median (interquartile range (IQR)) number of symptoms reported was 2 (1 to 4). The median (IQR) duration of symptoms prior to pre-registration was 4 (2 to 7) days. This duration was shorter for doctors at 1.5 (1 to 4) days. The most common symptom was headache, reported by 435 (54.44%) clients, followed by sore throat (361, 45.18%), cough (326, 40.80%) and myalgia (196, 24.53%). The frequency of other symptoms are shown in Table 2. Asymptomatic clients totalled 145 (18.15%).

**Table 2:** Frequency of symptoms amongst all clients pre-registering; and proportion of positive tests for SARS-CoV-2 according to the number of clients with a valid test result reporting the symptom.

| Symptom (N = 799) | Symptom frequency | Valid result | Positive result | Proportion (95% CI) |
| --- | --- | --- | --- | --- |
| Fever | 72 | 61 | 24 | 39.34 (28.06 ; 51.88) |
| Altered smell | 70 | 50 | 19 | 38.00 (25.86 ; 51.85) |
| Altered taste sensation | 92 | 65 | 23 | 35.38 (24.87 ; 47.52) |
| Chills | 75 | 60 | 18 | 30.00 (19.90 ; 42.51) |
| History of fever | 68 | 50 | 14 | 28.00 (16.67 ; 42.81) |
| General body weakness | 107 | 84 | 21 | 25.00 (16.47 ; 35.85) |
| Cough | 326 | 245 | 56 | 22.86 (18.04 ; 28.51) |
| Myalgia | 196 | 150 | 32 | 21.33 (15.53 ; 28.56) |
| Headache | 435 | 345 | 67 | 19.42 (15.59 ; 23.92) |
| Nausea and vomiting | 52 | 39 | 7 | 17.95 (8.10 ; 34.11) |
| Shortness of breath | 115 | 89 | 14 | 15.73 (9.61 ; 24.69) |
| Sore throat | 361 | 285 | 44 | 15.44 (11.71 ; 20.09) |
| Diarrhoea | 54 | 43 | 5 | 11.63 (5.07 ; 24.48) |
| Irritability | 23 | 12 | 1 | 8.33 (1.49 ; 35.38) |
| Asymptomatic | 145 | 99 | 12 | 12.12 (7.07 ; 20.00) |

There were 584 (73.09%) clients reporting contact with a confirmed or presumed COVID-19 case. This was self-reported and not verified. Of these, 472 (80.82%) were also symptomatic. Thirty three (4.13%) clients reported no symptoms or contact and had no indication to test according to pre-registration data, but 14 were still tested.

We found 105 positive results, representing 13.14% (95% CI: 10.97; 15.66) of clients using the pre-registration service and 17.77% (95% CI: 14.9; 21.06) of those with a valid positive or negative test result. This proportion was highest amongst nurses (53, 15.19%) compared to 6 (9.52%) doctors, 5 (8.62%) general assistants and 12 (8.00%) administrative staff. There were 486 (60.83%) negative tests, 8 (1.00%) indeterminate, 9 (1.13%) not run at the laboratory and 191 (23.90%) clients who pre-registered but weren’t tested.

Fever, altered smell, altered taste sensation and chills were the symptoms with the highest predictive value for SARS-CoV-2 infection as shown in Table 2. Diarrhoea and irritability had the lowest predictive value with < 10% of these clients testing positive. Twelve (8.28%) asymptomatic clients tested positive. This included 3 of 33 clients with no reported symptom or contact.

There was a strong association between a greater number of symptoms and a positive test (p = 0.0002). Clients with a positive test had a mean (SD) of 3.3 (0.4) symptoms compared to 2.5 (0.2) symptoms amongst clients with a negative test. We particularly noted an increased likelihood of a positive test in patients with 3 or 4+ symptoms (18.55% and 20.70% respectively) compared to 1 or 2 symptoms (8.57% and 6.75% respectively).

We also noted an increased likelihood of coronavirus infection in patients who had a 2 - 4 or 5 - 10 day duration of symptoms (17.70% and 16.95% respectively) compared to 0 - 1, 11 - 15 and 16+ days (9.45%, 11.11% and 7.41% respectively).

## Discussion

Streamlined staff testing is a critical component of a broader strategy to maximise the available frontline workforce in the face of the COVID-19 pandemic by decreasing absence.^[5]^ At our institution, 6 in 7 staff members tested negative for SARS-CoV-2 and were able to return to work earlier than they might otherwise have been able. The remainder were detected and isolated, preventing further spread.

Rapid and sustained uptake of the online pre-registration system shows that this format is acceptable to staff. It allows for ease of access, decreases waiting times and expedites test results through prioritisation of staff samples. During the period under review, unpublished departmental data showed that over 5 800 tests were performed at our TATC. Online pre-registration was therefore able to reduce inline clerical work of opening folders while clients waited by about 14%. Its use for clusters from other medical institutions and non-medical facilities demonstrates the potential for further expansion of the system.

Clients presenting via the pre-registration system were young with few comorbidities. It is unclear if this is reflective of the broader staff population at this academic institution. Alternatively it could indicate that risk assessment activities performed by the OHD had successfully shielded older, more at risk individuals from exposure; or that older staff members preferred not to use this hi-tech option. We were able to explain the large proportion of female clients in the study due to a large contingent of nursing staff testing.

Four staff groups were predominantly represented in the data: nurses, administrative staff, doctors and general assistants. This reflects staff with the most direct contact with patients, but other groups may be being missed. While doctors presented quickly, nurses, administrative staff and general assistants waited longer. This is a period of time staff remain at work and are potentially highly infectious.^[6]^ There is a need to identify barriers to testing and develop strategies to encourage earlier presentation.

A large proportion of staff pre-registered but never tested while a minority had laboratory issues but were not re-tested. This indicates a significant loss to follow up, but is in keeping with the broader experience at our facility precipitated by the massive testing loads as the pandemic progressed and is not specific to the pre-registration system. It is unclear if these potentially infected individuals continued to work without ever testing or continued to isolate for the full 14 days. Integration with a scaled up OHD will combat this in future and enhance surveillance and protection of staff and patients.^[7]^

We noted altered smell and altered taste sensation together with flu-like symptoms to be most predictive symptoms of SARS-CoV-2 infection. This reflects that findings noted globally in mildly symptomatic outpatients apply to our local population and shows that they can be incorporated into local guidelines.^[8]^

Over 8% of asymptomatic individuals, including several without a specific contact, tested positive for SARS-CoV-2. This has also been seen in other healthcare worker screening programmes.^[9,10]^ Many such asymptomatic and pre-symptomatic individuals may remain undetected as a vector for nosocomial spread to other staff and vulnerable patients.^[11]^ In a system reliant on self-presentation, asymptomatic and oligo-symptomatic individuals will inevitably be missed. Changes to the provincial community screening and testing strategy advising most symptomatic individuals to self-isolate without testing have significantly reduced the number of tests being performed at the TATC and an alternative surveillance system testing all staff at intervals may now be feasible.^[12]^

It remains an unanswered question whether future staff testing and isolation strategies can exclude sufficiently low-risk symptomatic presentations allowing staff to remain at work with suitable precautions in place while awaiting a result. Such a strategy could reduce unnecessary isolation and workforce depletion. In this study, no symptom could completely eliminate the risk, while asymptomatic infection is also shown. We found it preferable to focus on rapid turnaround of results.

Limitations in this study are its retrospective nature and the reliance on client self-reported clinical information.

## Conclusion

Staff coronavirus testing using an online pre-registration form is a viable and acceptable strategy. While it does require a degree of technical know-how to set up, it can be inexpensively implemented using free or low cost third-party services. Benefits include streamlined testing, reduced clerical workload and errors, ease of data capturing and decreased risk of cross-infection. This strategy can replace or complement existing on-site systems.

The primary challenges that need to be overcome for a successful staff testing programme are following up non-attendance and ensuring a rapid turnaround of test results. Institutions also need to ensure that staff test timeously.

Hospital staff remain at high risk for SARS-CoV-2 infection and, once infected can become vectors for institutional outbreaks. Current guidelines provide advice on when to test and isolate staff who are symptomatic or have high- or low-risk contacts. These should be continuously updated as we learn more about the virus taking a balanced view to protect staff and maintain the available workforce. Staff testing must also be seen as one component of a larger strategy aimed at protecting staff, such as wearing masks and face shields, regular hand washing, buddy screening, physical distancing, availability of PPE and special dispensation for COVID-19-related leave.

## Data Availability

Raw data were generated at Tygerberg Hospital. Derived data supporting the findings of this study are available from the corresponding author on request.

## Declarations

## Acknowledgements

The staff of the TBH COVID-19 Unit for their selflessness and dedication in caring for our patients, and in particular the clerical staff who open a prodigious number of folders every day.

## Author Contributions

MSM: concept, literature review, data collection, data analysis, manuscript completion; AP, MAP, SS, AM: literature review, manuscript completion; LA, RC, TB: data collection, manuscript completion.

## Data availability statement

Raw data were generated at Tygerberg Hospital. Derived data supporting the findings of this study are available from the corresponding author MSM on request.

## Disclaimer

The views expressed in this article are those of the author and not an official position of the institution or journal.

